# Characterisation of post-fundoplication gastric dysfunction using Gastric Alimetry

**DOI:** 10.1101/2023.11.05.23297357

**Authors:** William Xu, Tim Wang, Daphne Foong, Gabe Schamberg, Nicholas Evennett, Grant Beban, Armen Gharibans, Stefan Calder, Charlotte Daker, Vincent Ho, Greg O’Grady

## Abstract

**Background:** Adverse gastric symptoms persist in up to 20% of fundoplication surgeries completed for gastroesophageal reflux disease, causing significant morbidity, and driving the need for revisional procedures. Non-invasive techniques to assess the mechanisms of persistent postoperative symptoms are lacking. We aimed to investigate gastric myoelectrical abnormalities and symptoms in patients after fundoplication using a novel non-invasive body surface gastric mapping (BSGM) device.

**Methods:** Patients with previous fundoplication surgery and ongoing significant gastroduodenal symptoms, and matched controls were included. BSGM using Gastric Alimetry (Alimetry, New Zealand) was employed, consisting of a high resolution 64-channel array, validated symptom-logging App, and wearable reader.

**Results:** 16 patients with significant chronic symptoms post-fundoplication were recruited, with 16 matched controls. Overall, 6/16 (37.5%) patients showed significant spectral abnormalities defined by unstable gastric myoelectrical activity (n = 2), abnormally high gastric frequencies (n = 3) or high gastric amplitudes (n = 1). Those with spectral abnormalities had higher Patient Assessment of Upper Gastrointestinal Disorders-Symptom Severity Index scores (3.2 [2.8 to 3.6] vs 2.3 [2.2 to 2.8]; p =0.024). 7/16 patients (43.8%) had Gastric Alimetry tests suggestive of gut-brain axis contributions, and without myoelectrical dysfunction. Increasing Principal Gastric Frequency deviation, and decreasing Rhythm Index were associated with symptom severity (r>0.40, p<0.05).

**Conclusion:** A significant number of patients with persistent post-fundoplication symptoms display abnormal gastric function on Gastric Alimetry testing, which correlates with symptom severity. These findings advance the pathophysiological understanding of post-fundoplication disorders which may inform diagnosis and patient selection for medical therapy and revisional surgery.

## Background

The global prevalence of gastro-oesophageal reflux disease is high and increasing.^1^ Anti-reflux surgeries via fundoplication are commonly performed for patients with disease refractory to proton pump inhibitors (PPIs), typically in those who wish to discontinue medical therapy, or have concurrent large hiatal hernias or refractory esophagitis.^2^

While the long-term efficacy of fundoplication surgery for chronic GERD symptoms is comparable to medical therapy, significant postoperative adverse effects can occur, with a minority of patients developing chronic gastroduodenal symptoms. However, only around 40% of patients with persistent symptoms have gastroesophageal reflux confirmed on oesophageal function tests,^3^ with the remaining patients often developing symptoms akin to post-fundoplication dyspepsia or gastroparesis.

Post-fundoplication dyspepsia is characterised by bloating, and early satiation,^4,5^ with some also developing more debilitating abdominal pain and nausea.^6,7^ In most, these symptoms self-resolve within 12 months, but approximately 5-16% patients develop chronic symptoms that impact quality of life ^6,8,9^

The pathogenesis of post-fundoplication symptoms is multifactorial, with post-surgical gastric dysmotility, accelerated or delayed gastric emptying, impaired gastric accommodation, altered lower oesophageal sphincter pressure, or vagal nerve injury at time of surgery, all postulated as contributing mechanisms.^5,8,10,11^ It is clinically challenging to distinguish the underlying cause of post-fundoplication symptoms, and tests such as endoscopy, pH-impedance, and manometry are often non-contributory, as well as being moderately invasive and resource intensive. Consequently, management pathways of such patients remain suboptimal and unclear.^7^ A safe, accurate, and non-invasive test is needed to reliably assess gastric function to help inform correct therapy.

Body surface gastric mapping (BSGM; also termed high-resolution electrogastrography) using Gastric Alimetry^Ⓡ^(Alimetry, New Zealand) is a novel diagnostic technique employing a dense array of electrodes at the epigastrium, which has been extensively validated for evaluation of gastric electrophysiological function.^12–14^ We hypothesised these methods would provide novel insight into the pathophysiology of post-fundoplication symptoms, offering an improved mechanistic basis for treatment. The aim of this study was therefore to characterise gastric myoelectrical abnormalities in patients with persistent symptoms after fundoplication surgery using the Gastric Alimetry system.

## Methods

Ethical approval for reporting was obtained from the Auckland Health Ethics Research Ethics Committee and Western Sydney Human Research Ethics Committee (ref: AH1130, H15157).

Consecutive adult patients who had a previous fundoplication with ongoing upper gastrointestinal symptoms referred for Gastric Alimetry evaluation at Auckland New Zealand, or Western Sydney, Australia were approached for inclusion. Patients were excluded if they had undergone gastric bypass surgery as revisional surgeries, had an active gastrointestinal malignancy, an active neurogenic or endocrine disorder affecting gastric motility (multiple sclerosis, scleroderma, Parkinson’s disease, hyperthyroidism), a current pregnancy, or cognitive impairment. No restriction was put on the timing of previous fundoplication surgery. Routine Gastric Alimetry test exclusions were also applied (BMI>35, fragile or damaged epigastric skin, unable to sit in a still position, or major allergy to adhesives).^12^

### Gastric Alimetry procedure

All patients underwent a standard Gastric Alimetry test for evaluation of gut function, as described in detail elsewhere.^15,16^ In brief, patients withheld GI motility affecting medications such as prokinetics and fasted for 8 hours prior, before undergoing a 30-minute baseline recording, followed by a standardised meal consisting of Ensure (232 kcal, 250 mL; Abbott Nutrition, IL, USA) and an oatmeal energy bar (250 kcal, 5 g fat, 45 g carbohydrate, 10 g protein, 7 g fibre; Clif Bar & Company, CA, USA), and a 4-hr postprandial recording. Patients were encouraged to eat as much of the meal as possible in order to stimulate symptoms.^15^

The Gastric Alimetry device consists of a high-resolution stretchable electrode array (8 × 8 electrodes; 20 mm spacing; 196 cm2), a wearable Reader, validated iOS app for symptom logging and a cloud-based reporting platform.^14^ (**Figure 1**) Analysis of tests involves automated artefact rejection followed by the assessment of four validated test metrics: Gastric Alimetry Rhythm Index^TM^ (GA-RI; a measure of gastric rhythm stability), the Principal Gastric Frequency, BMI-adjusted amplitude, and the Fed:Fasted amplitude ratio. Details of these metrics and the establishment of normative intervals are described in detail elsewhere.^17^ Additionally, given the transient nature of Principal Gastric Frequency abnormalities seen in this cohort, maximum absolute Principal Gastric Frequency Deviation (the maximum absolute difference of Principal Gastric Frequency from 3cpm for any given postprandial hour) was calculated to quantify the magnitude of transient gastric dysrhythmias.

**Figure 1:**
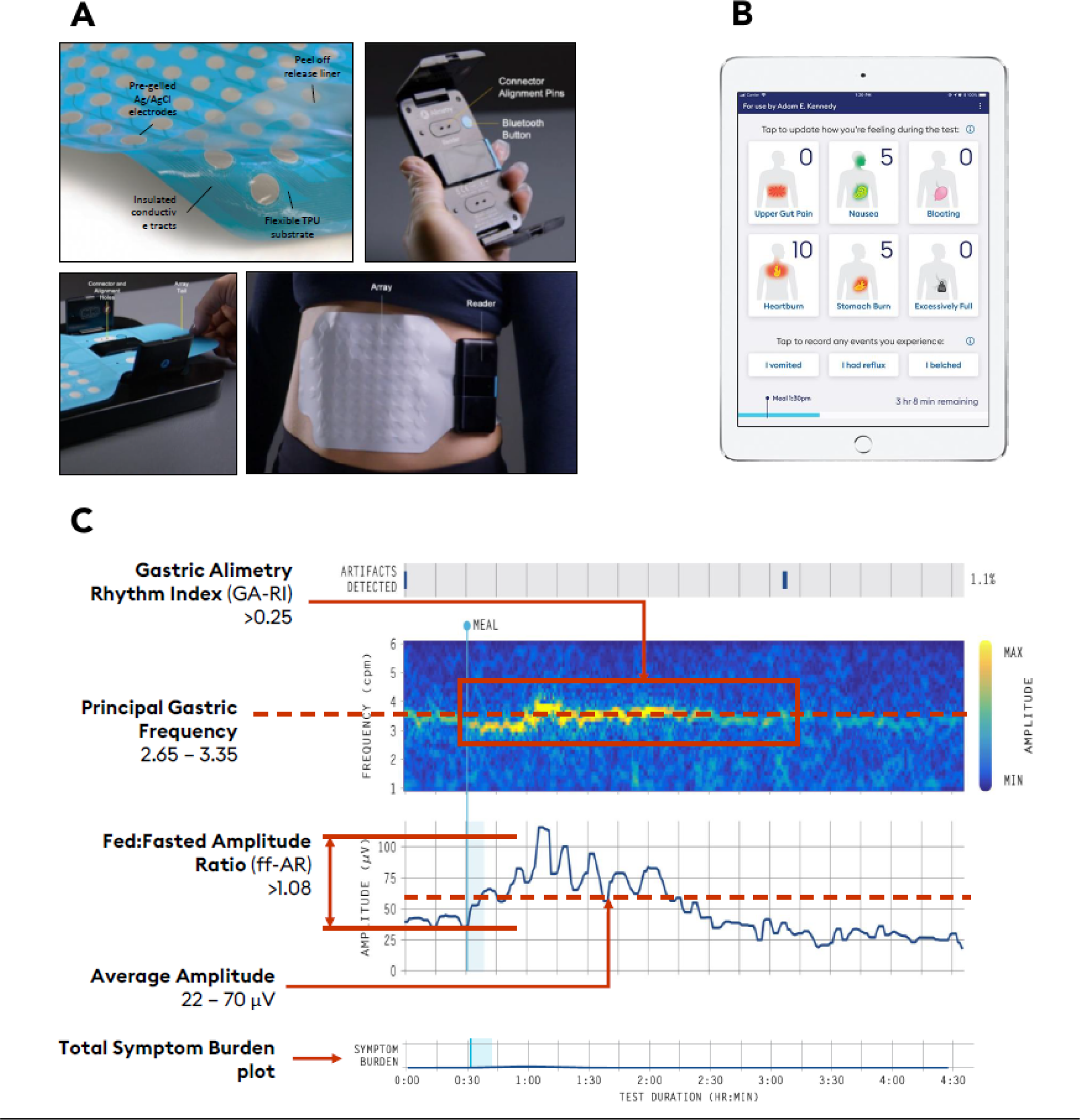
The Gastric Alimetry System. (A) The Gastric Alimetry™ BSGM system (B) The Gastric Alimetry App™ used for setup, data transfer, and symptom data tracking. (C) An example of a normal Gastric Alimetry test with reference ranges of Gastric Alimetry Spectral Outputs and the Total Symptom Burden plot

Based on the above Gastric Alimetry metrics, patients were subsequently evaluated and classified to the following established phenotypes:^12^

1. *Gastric neuromuscular dysfunction:* Gastric rhythm disorder or low amplitude, indicated by Gastric Alimetry Rhythm Index (GA-RI) <0.25, and/or BMI-adjusted amplitude <22 μV.^18^
2. *High sustained BMI-adjusted amplitude*: >70 μV suggesting possible gastric outlet resistance.^18,19^
3. *High frequency phenotype:* >3.35cpm suggesting possible vagal nerve dysfunction.^16,20,21^

During testing, patients simultaneously logged their gastroduodenal symptom profiles in the validated Gastric Alimetry App.^22^ For patients with normal spectral metrics (i.e. frequency, rhythm and amplitude), symptom profiles were analysed in relation to onset of gastric activity and evaluated according to the following additional established phenotypes.^23^

1. *Amplitude-dependent symptoms:* The onset of symptoms is related to the timing of gastric activity *Sensorimotor:* symptoms relating directly with gastric amplitude (r>0.5), suggesting a potential hypersensitivity / accommodation disorder.^23,24^

*Post-gastric*: symptoms trending upward late in postprandial period; after the gastric meal response had peaked, indicating a likely more distal pathology (e.g., small bowel dysmotility or other disorders).^25^

1. ii) *Amplitude-independent symptoms*: symptoms continuous/do not relate to gastric activity, suggesting possible gut-brain axis disorders.^23^

### Symptom assessment

Baseline symptom-severity and quality-of-life was completed with the Patient Assessment of Upper Gastrointestinal Disorders-Symptom Severity Index (PAGI-SYM), Gastroparesis Cardinal Symptom Index (GCSI), and Patient Assessment of Upper Gastrointestinal Disorders-Quality of Life (PAGI-QOL) instruments.^26,27^ Health psychology assessment were completed using the Patient Health Questionnaire-2 (PHQ-2) questionnaire.^28^ The Rome IV criteria for chronic nausea and vomiting syndrome or functional dyspepsia were used to determine a threshold for a clinically substantial chronic upper gastrointestinal symptom burden.^29^ Total Symptom Burden was calculated, as the sum of each participant’s mean postprandial symptom scores including early satiation.^22^

### Patient matching

Post-fundoplication participants were matched to a database of controls (110 subjects ≥18 years) in a 1:1 ratio using the nearest neighbour based on age, sex, and body mass index (BMI), with the matchit package.^30^ Control subjects were excluded if they had active gastrointestinal symptoms, met Rome IV criteria, were taking medications altering gastrointestinal motility, or consumed regular cannabis. Tests with >50% of the duration marked as artefacts were also excluded from the analysis, per the Gastric Alimetry Instructions for Use.

### Statistical analysis

Data are reported as the mean ± standard deviation (SD) or median and interquartile range (IQR). Comparisons were made using one-way analysis of variance (ANOVA) tests with further pairwise comparisons corrected for by the Tukey post-hoc test. The chi-squared test was used for categorical variables. Continuous independent variables were compared using the Mann-Whitney U test. Associations between demographic data, BSGM metrics, patient symptom symptoms and quality of life were assessed using Pearson correlation with Benjamini-Hochberg’s corrections to minimise false discovery rate. A statistical significance threshold P<0.05 was used. All analyses were performed in R version 4.2.0 (R Foundation for Statistical Computing, Vienna, Austria)

## Results

Gastric Alimetry studies were completed in 16 patients with persistent symptoms after fundoplication (median age 34.5 years, (range 22-65); 6 females (37.5%), mean BMI 26.6 ± 5.9), which were successfully matched by age, sex, and BMI to 16 healthy controls (**Table 1**). Median time of Gastric Alimetry testing was 5 years after index fundoplication surgery (range 0 to 19). A total of 6 patients (37.5%) had revisional surgery prior to Gastric Alimetry, and 2 additional patients (12.5%) had surgical reversal of fundoplication. All patients had index surgery for symptomatic gastroesophageal reflux and all patients had concurrent hiatal hernias. Based on the Rome classification for chronic nausea and vomiting syndrome (CNVS) and functional dyspepsia (FD), a total of 13 patients met criteria for both CNVS and FD and 3 patients met criteria for FD only (**Table 1**).

**Table 1:** Demographic and surgical details. Data are presented as mean ± SD, median (interquartile range) or number (%).

| Variable |  | Control | Spectral-N | Spectral-Abn | Total | p |
| --- | --- | --- | --- | --- | --- | --- |
| **Total N (%)** |  | **16 (50.0)** | **10 (31.2)** | **6 (18.8)** | **32** |  |
| Age (years) | | 39.0 ± 16.5 | 42.1 ± 14.9 | 36.3 ± 12.6 | 39.5 (15.0) | 0.758 |
| Sex (female) |  | 9 (56.2) | 3 (30.0) | 3 (50.0) | 15 (46.9) | 0.412 |
| BMI (kg/m<sup>2</sup>) | | 26.0 ± 4.0 | 27.2 ± 5.8 | 25.6 ± 6.5 | 26.3 ± 5.0 | 0.787 |
| Irritable bowel syndrome |  | 1 (6.2) | 2 (20.0) | 2 (33.3) | 5 (15.6) | 0.184 |
| Type 2 Diabetes |  | 0 (0.0) | 1 (10.0) |  | 1 (3.1) | 0.5 |
| Depression |  | 0 (0.0) | 4 (40.0) | 2 (33.3) | 6 (18.8) | 0.014 |
| Anxiety |  | 0 (0.0) | 5 (50.0) | 2 (33.3) | 7 (21.9) | 0.004 |
| PHQ-2 score |  | 0.0 (0.0 to 0.2) | 2.0 (0.5 to 2.0) | 1.0 (0.0 to 5.0) | 0.0 (0.0 to 2.0) | 0.043 |
| ROME classification | CNVS + FD | 0 (0.0) | 7 (70.0) | 6 (100.0) | 13 (40.6) | <0.001 |
|  | FD only | 0 (0.0) | 3 (30.0) | 0 (0.0) | 3 (9.4) |  |
| Years since index surgery |  | - | 4.5 (2.0 to 5.8) | 11.5 (5.0 to 16.5) | 5.0 (3.5 to 10.0) | 0.064 |
| Type of index fundoplication | Partial anterior | - | 3 (30.0) | 1 (16.7) | 4 (25.0) | 0.453 |
|  | Partial posterior | - | 2 (20.0) | 0 (0.0) | 2 (12.5) |  |
|  | Total posterior | - | 5 (50.0) | 5 (83.3) | 10 (62.5) |  |
| Revisional surgery |  | - | 3 (30.0) | 3 (50.0) | 6 (37.5) | 0.607 |
| Fundoplication reversal surgery |  | - | 2 (20.0) | 0 (0.0) | 2 (12.5) | 0.500 |
| Gastric Emptying Results | Delayed | - | 6 (60.0) | 4 (66.7) | 10 (62.5) | 1.000 |
|  | Missing | - | 4 (40.0) | 2 (33.3) | 6 (37.5) |  |

### Gastric Alimetry classification of patients

Of 16 cases, 6 patients had abnormal Gastric Alimetry tests (37.5%), of which 2 patients showed gastric neuromuscular dysfunction characterised by low gastric rhythm stability (12.5%), 3 showed transient abnormally high gastric frequencies (18.75%), and one demonstrated abnormally high gastric amplitudes (6.25%; **Figure 2**). Averaged spectrograms of each abnormal spectral phenotype group and the control group are displayed in **Figure 2**. Full Gastric Alimetry classifications of each patient are described in **Table S1.** Individual Gastric Alimetry spectral plots are presented in **Figures S1** and **S2**.

**Figure 2:**
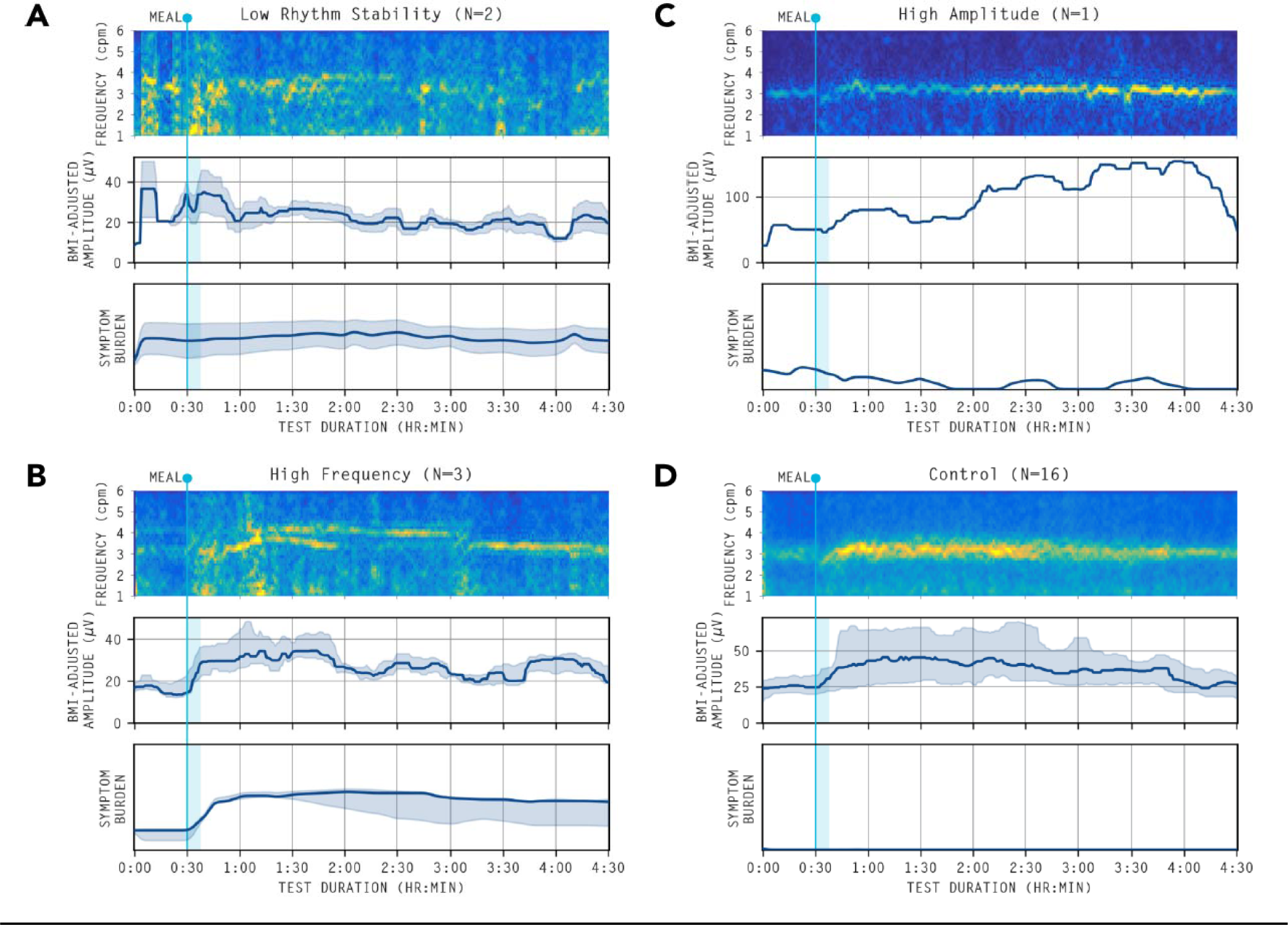
Averaged spectrograms and amplitude curves of abnormal spectrograms. Averaged spectrograms of the abnormal spectral analyses. (A) Gastric neuromuscular dysfunction Gastric rhythm disorder or low amplitude, indicated by Gastric Alimetry Rhythm Index (GA-RI) <0.25, and/or BMI-adjusted amplitude <22 μV.^18^ (B) High frequency phenotype: >3.35cpm suggesting possible vagal nerve dysfunction.^16^ (C) High sustained BMI-adjusted amplitude: >70 μV suggesting possible gastric outlet resistance.^20^ (D) Healthy controls.

Of note, two out of three patients with abnormally high gastric frequencies (66.6%) had presumed vagal nerve injury documented in procedure notes. In case 14, the patient had revisional anterior fundoplication surgery after an initial Nissen fundoplication. On revisional surgery, the anterior vagus nerve was identified and preserved but the posterior vagus nerve was unable to be visualised. Dense adhesions were present and the posterior vagus nerve was assumed to have been surgically injured. In case 15, the patient had a Nissen fundoplication with a possible vagal nerve injury documented at index procedure.

The remaining 10 post-fundoplication patients (62.5%) had normal Gastric Alimetry spectrograms (**Figure S3**), comprising 6 patients (37.5%) with *amplitude-independent* symptom profiles, and 4 patients (25%) with *amplitude-dependent* symptom profiles. Two patients (12.5%) had sensorimotor phenotypes, defined by a correlation between symptom severity and gastric amplitude (r>0.5). None of these 10 patients had vagal nerve injuries documented in their procedural notes.

### Group-wise differences between normal and abnormal Gastric Alimetry

Gastric Alimetry metrics and symptom severity between controls patient groups are reported in **Table 2** and **Figure 3**.

**Figure 3:**
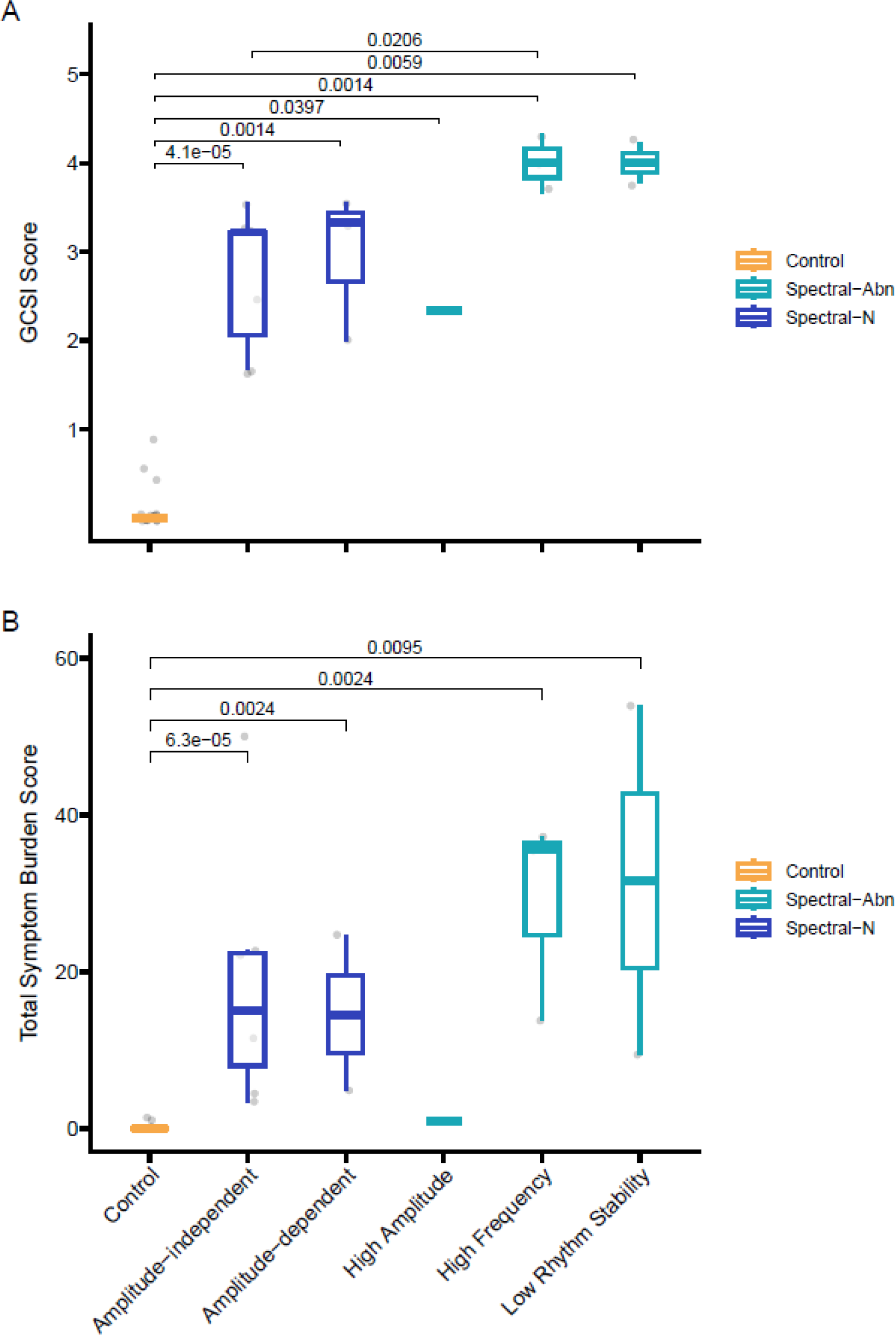
GCSI and Total Symptom Burden by Gastric Alimetry Phenotype. Symptom burden by Gastric Alimetry phenotype when measured by (A) Gastroparesis Cardinal Symptom Index (GCSI) and (B) Total Symptom Burden as measured simultaneously during the Gastric Alimetry Test. Groups compared with the Wilcoxon Rank Sum test with corresponding significant p-values displayed (p<0.05). Spectral-Abn: Abnormal Gastric Alimetry Spectral analysis. Spectral-N: Normal Gastric Alimetry Spectral analysis.

**Table 2:** Metrics and symptoms between columns. Principal Gastric Frequency Deviation is calculated as the highest absolute deviation from 3cpm for any 1-hour period postprandially.

| Variable | Control | Spectral-N | Spectral-Abn | Total | Control vs Spectral-Abn p-value | Control vs Spectral-N p-value | Spectral-N vs Spectral-Abn p-value |
| --- | --- | --- | --- | --- | --- | --- | --- |
| **Total N (%)** | **16 (50.0)** | **10 (31.2)** | **6 (18.8)** | **32** |  |  |  |
| Adjusted Amplitude (µV) | 37.9 (29.2 to 51.7) | 35.9 (29.8 to 41.1) | 29.0 (26.0 to 33.5) | 34.5 (29.2 to 44.3) | 0.925 | 0.716 | 0.963 |
| Fed Fasted - Amplitude Ratio | 2.0 (1.2 to 2.4) | 1.8 (1.5 to 2.2) | 1.9 (1.3 to 2.1) | 2.0 (1.4 to 2.2) | 0.518 | 0.868 | 0.806 |
| Principal Gastric Frequency (cpm) | 3.1 (2.9 to 3.2) | 2.9 (2.8 to 3.0) | 3.3 (3.2 to 3.5) | 3.1 (2.9 to 3.2) | **0.035** | 0.141 | **0.001** |
| Principal Gastric Frequency Deviation (from 3cpm) | 0.3 (0.2 to 0.4) | 0.2 (0.2 to 0.4) | 0.6 (0.3 to 1.0) | 0.3 (0.2 to 0.5) | **0.008** | 0.868 | **0.005** |
| Gastric Alimetry Rhythm Index | 0.5 (0.4 to 0.8) | 0.5 (0.4 to 0.6) | 0.3 (0.2 to 0.4) | 0.5 (0.4 to 0.6) | 0.218 | 0.704 | 0.607 |
| Meal Completion (%) | 100.0 (100.0 to 100.0) | 100.0 (92.5 to 100.0) | 85.0 (62.5 to 100.0) | 100.0 (97.5 to 100.0) | 0.251 | 0.893 | 0.490 |
| Percent Artefact (%) | 9.0 ± 6.7 | 16.7 ± 7.7 | 18.2 ± 14.6 | 13.1 ± 9.6 | 0.096 | 0.097 | 0.945 |
| PAGI-SYM Score | 0.0 (0.0 to 0.4) | 2.3 (2.2 to 2.8) | 3.2 (2.8 to 3.6) | 1.0 (0.0 to 2.6) | **<0.001** | **<0.001** | **0.021** |
| GCSI Score | 0.0 (0.0 to 0.0) | 3.2 (2.1 to 3.3) | 3.9 (3.7 to 4.2) | 1.3 (0.0 to 3.2) | **<0.001** | **<0.001** | **0.008** |
| PAGI-QoL Score | 0.2 (0.0 to 0.3) | 2.3 (1.9 to 3.2) | 3.4 (2.0 to 3.6) | 0.8 (0.2 to 2.6) | **<0.001** | **<0.001** | 0.678 |
| Total Symptom Burden | 0.0 (0.0 to 0.0) | 14.8 (6.5 to 22.5) | 24.6 (10.5 to 36.8) | 1.2 (0.0 to 14.6) | **<0.001** | **0.002** | 0.393 |

#### Rhythm stability and gastric frequency

There was no overall group-wise difference in Gastric Alimetry metrics between post-fundoplication patients and controls (**Table S2**). However, post-fundoplication patients with abnormal Gastric Alimetry spectral analyses showed higher overall Principal Gastric Frequencies compared to patients without spectral abnormalities (3.3 [IQR: 3.2 to 3.5] abnormal spectral vs 2.9 [2.8 to 3.0] normal spectral, p=0.001, **Table 2**) and compared to controls (3.3 [IQR: 3.2 to 3.5] vs 3.1 [2.9 to 3.2] controls, p=0.035, **Table 2**). These patients also displayed a higher maximum Principal Gastric Frequency deviation compared with patients with normal spectral analysis (0.6 [IQR: 0.3 to 1.0] vs 0.2 [0.2 to 0.4], p = 0.008, **Table 2**) and with controls (0.6 [IQR: 0.3 to 1.0] vs 0.3 [0.2 to 0.4], p = 0.005, **Table 2**).

#### Symptom severity

Post-fundoplication patients with spectral abnormalities had higher PAGI-SYM scores compared to patients without spectral abnormalities (3.2 [2.8 to 3.6] abnormal spectral vs 2.3 [2.2 to 2.8] normal spectral, p =0.024, **Table 2**, **Figure 3**) and compared to controls (3.2 [2.8 to 3.6] abnormal spectral vs 0 [0 to 0.4] controls, p<0.001). The severity of nausea, bloating, upper abdominal pain, stomach burn, heartburn, early satiation and excessive fullness symptoms between those with spectral abnormalities and those without are presented as radial plots in **Figure S4** and **Table S3.**

### Clinical associations between Gastric Alimetry metrics and patient factors

Correlations between patient symptoms and Gastric Alimetry metrics are presented in **Figure 4**. On univariate analysis there was a significant correlation between the maximum absolute Principal Gastric Frequency deviation and Total Symptom Burden severity (r=0.41, p = 0.021; **Figure 4B**). A lower Rhythm Index (GA-RI) was also associated with an increasing Total Symptom Burden severity (r=-0.42, p = 0.016; **Figure 4C**). No associations were found between Total Symptom Burden severity and BMI-adjusted amplitude, fed-fasted amplitude ratio, or years since index surgery (**Figure 4D-F**).

**Figure 4:**
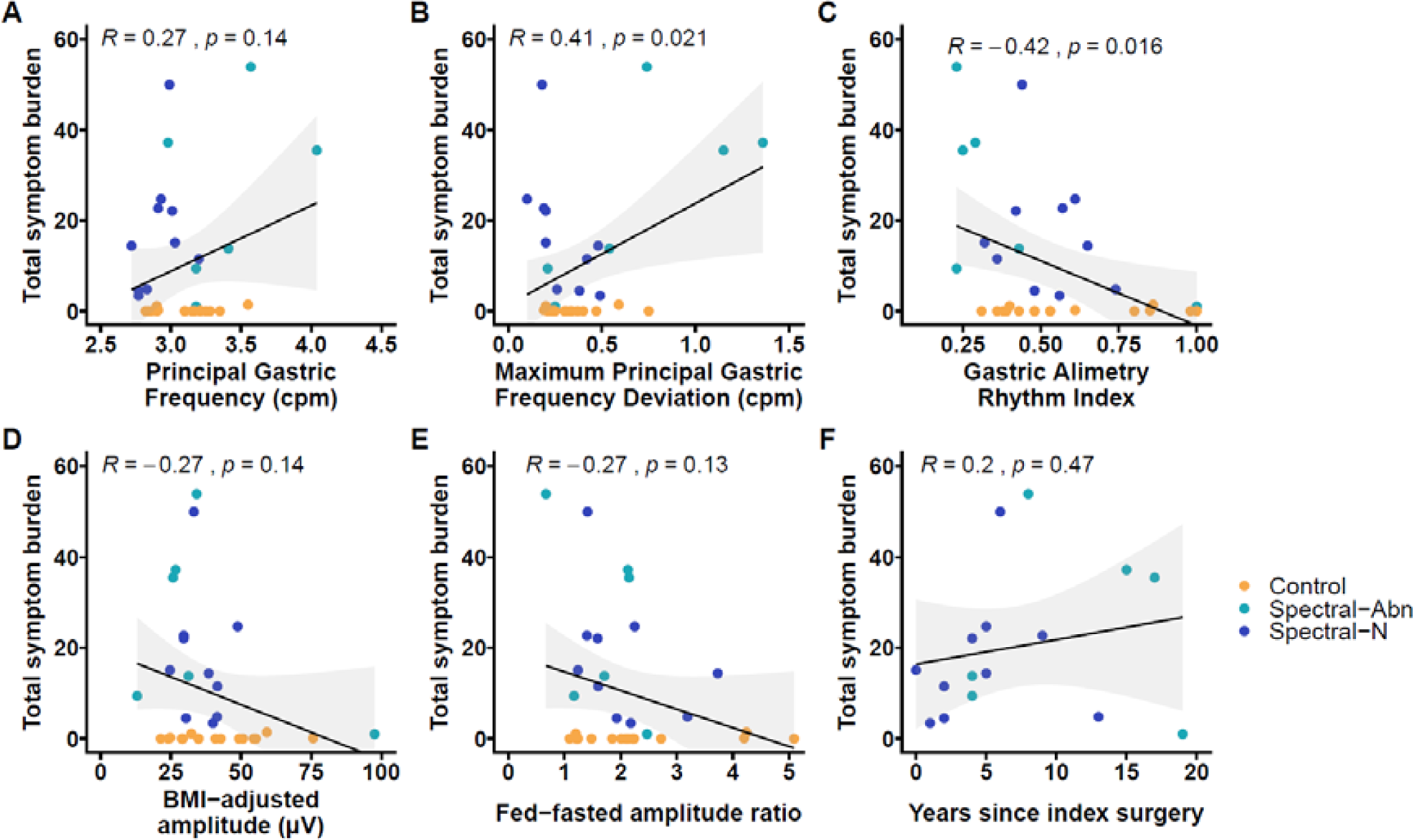
Scatter plots of association between symptom burden and Gastric Alimety metrics. Scatter plots displaying the association between Total Symptom Burden as measured simultaneously during the Gastric Alimetry Test and (A) Principal Gastric Frequency (cycles per minute) (B) maximum absolute deviation of Principal Gastric Frequency from 3cpm for any 1 hour postprandial period of the test (C) Gastric Alimetry Rhythm Index (D) BMI-adjusted amplitude µV (E) Fed-fasted amplitude ratio and (F) Years since index fundoplication surgery

Adjusted correlations post-Benjamini Hochberg’s corrections between symptoms, surgical history, and BSGM metrics are displayed in **Figure 5**. Increasing Principal Gastric Frequency deviation, and decreasing GA-RI were each significantly associated with severity of symptoms including fullness, bloating, early satiation, and total symptom burden on whole cohort analysis (**Figure 5A**, n=32, r>0.40, p<0.05).

**Figure 5:**
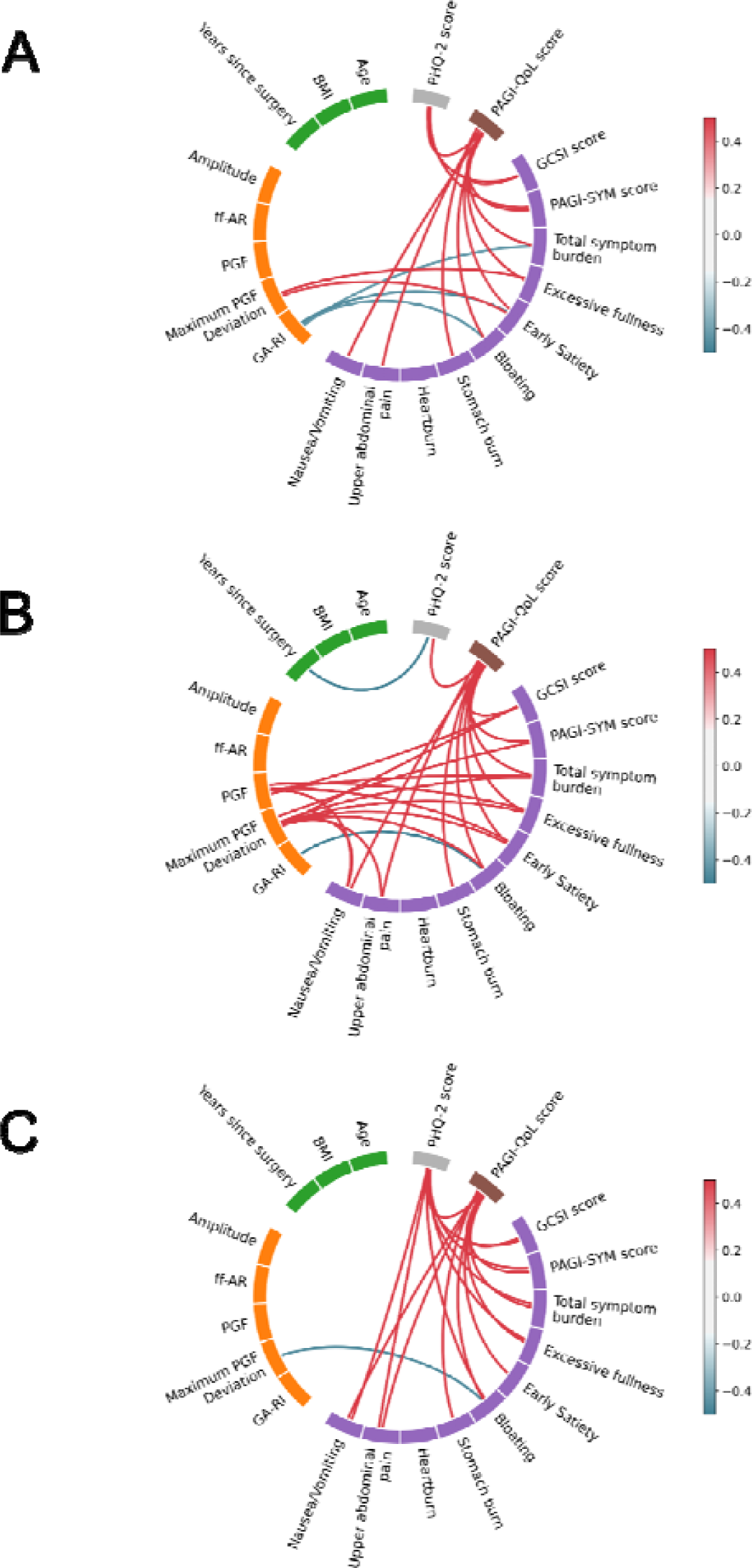
Correlation wheel plots. Correlations between variables, adjusted for multiple comparisons using the Benjamini-Hochberg correction. (A) All patients and controls (n=32), (B) patients with spectral abnormalities and controls (n=22), (C) patients without spectral abnormalities and controls (n=26). Variables are grouped into categories:patient demographic factors, Gastric Alimetry metrics, symptom scores, quality of life, and psychological comorbidity. Correlations within categories are not shown. Only statistically significant correlations are shown (adjusted p<0.05)

On subgroup analysis of patients with abnormal spectral analyses, Gastric Alimetry test metrics, but not depression scale scores, correlated with symptom severity when compared with controls (**Figure 5B**, n=22, r>0.45, p<0.05). In particular, maximum absolute Principal Gastric Frequency deviation was strongly correlated with upper gut pain (r=0.63, p = 0.006), nausea (r = 0.66, p = 0.001), bloating (r=0.71, p <0.001), early satiation (r=0.81, p <0.001), excessive fullness (r=0.83, p <0.001), Total Symptom Burden (r=0.75, p <0.001), PAGI-SYM score (r=0.60, p=0.01), and GCSI score (r=0.61, p =0.009, **Figure 5B**).

Conversely, on subgroup analysis of patients with normal BSGM spectral analyses and controls, depression scores measured by the PHQ-2 score, correlated with symptom severity of nausea, abdominal pain, bloating, early satiation, excessive fullness, total symptom burden and PAGI-SYM/GCSI scores (**Figure 5C**, n=26, all r>0.40, p<0.05).

### Gastric emptying

In total, 8 out of 16 patients (50.0%) had documented evidence of delayed gastric emptying on post-fundoplication gastric emptying studies. Another 2 patients had retained food noted on gastroscopy despite a prolonged fast of 6 hours or more and hence were classified as delayed gastric emptying based on clinical grounds. Delayed emptying was observed in patients both with and without Gastric Alimetry spectral abnormalities (p=1.00; **Table 1**).

## Discussion

This study applied BSGM to evaluate the mechanisms of persistent post-fundoplication symptoms. Approximately 40% of patients in our cohort displayed abnormal gastric myoelectrical activity, characterised by persistently high gastric frequencies, unstable gastric rhythms, or abnormally high gastric amplitudes. A further 44% of patients had symptoms that did not correlate with gastric activity but did correlate with depression scores, suggesting gut-brain axis contributions. Gastric Alimetry was therefore able to distinguish patients with gastric neuromuscular dysfunction after fundoplication surgery from those with normal gastric activity, providing novel targets to inform diagnosis and therapy.

Abnormally low Gastric Alimetry Rhythm Index, or abnormally high Principal Gastric Frequency, were directly correlated with symptom severity in post-fundoplication patients. Of particular significance, 19% of post-fundoplication patients, all with a substantial gastroduodenal symptom burden, displayed a markedly elevated Principal Gastric Frequency. The vagus nerve is important in regulating ICC frequency,^31^ with stable high gastric frequencies demonstrated after vagotomy in previous studies.^32,33^ In addition, abnormally high gastric frequency has also recently been observed in patients with longstanding type 1 diabetes and peripheral neuropathy, likely on the basis of co-existent autonomic neuropathy.^16^ Therefore, high frequency gastric activity appears to be a general indicator of vagal nerve dysfunction or injury, although its sensitivity as a biomarker for this pathology requires further research.

Vagal nerve injury has long been implicated as an important pathogenic mechanism in post-fundoplication symptoms. The vagus nerve has significant roles in gastric physiology including mediating gastric accommodation,^5,34^ regulating liquid and solid gastric emptying,^35^ and regulation of ICC frequency.^31^ One historic case series specified a 4% vagal nerve injury rate in laparoscopic fundoplication, in which the majority of these patients subsequently developed symptoms of gastroparesis.^36^ Rates of vagal injury in other retrospective series have been reported as high as 20%, and were associated with poorer outcomes.^37^ Richards et al. also demonstrated that Nissen fundoplication was associated with high gastric frequencies postoperatively in children.^38^ Studies on long term outcome of gastric vagotomy report dyspeptic symptoms in 18% of patients.^21,39^ Two out of three patients with Gastric Alimetry findings of high gastric frequency in this cohort had high clinical suspicion of damaged vagus nerves at time of surgery, furthering the hypothesis that high gastric frequency is a useful clinical biomarker of vagal dysfunction or injury.

The other significant phenotype identified in this cohort is the group displaying signs of gastric neuromuscular dysfunction. Reduced GA-RI is considered to be reflective of gastric neuromuscular dysfunction secondary to depleted interstitial cells of Cajal networks or sustained gastric dysrhythmia, a finding demonstrated in patients with diabetic or idiopathic gastroparesis.^40,41^ ICC depletion leads to disordered gastric slow-wave conduction and contractile dysfunction,^31,42^ and is reflected by weak irregular gastric myoelectrical activity such as in patients with nausea and vomiting disorders.^15^ It is unclear how ICC depletion occurs mechanistically after fundoplication surgery, although there may be a survival dependency between ICC and vagal afferent nerve endings, indicating that vagotomy could again be a contributing factor.^43^ Further studies correlating BSGM data with histological data in post-fundoplication patients would be of interest.

The variety of other patient subgroups found in this study supports the view that chronic post-fundoplication symptoms arise from heterogeneous mechanisms. We identified two patients (12.5%) with normal gastric myoelectrical activity but a strong correlation between meal ingestion, gastric amplitude, and symptom severity, classified as a ‘sensorimotor’ symptom profile according to Gastric Alimetry criteria.^25^ This finding has previously been postulated to reflect a gastric accommodation disorder or visceral hypersensitivity.^23,25^ Significantly impaired gastric accommodation impairment has also been previously demonstrated in those with post-fundoplication dyspepsia symptoms.^5^ The impairment of gastric accommodation is likely multifactorial, with contributions from anatomical distortion of the fundus, surgical manipulation of the gastro-oesophageal junction, and vagal dysfunction.^5^

Furthermore, 6 patients (37.5%) had normal Gastric Alimetry spectrograms and amplitude-independent symptom profiles, with significant associations between PHQ-2 depression scores and symptom severity. In medical cohorts without prior surgery, this phenotype has been suggested to indicate dominant pathophysiological contributions from the gut-brain axis,^15^ however, this is a novel finding in post-surgical patients that now requires further research.

Treatment options for post-fundoplication symptoms are not well established, but phenotyping patients into mechanistic subgroups using Gastric Alimetry may provide a critical advance in guiding treatment decisions. Patients with signs of gastric neuromuscular dysfunction, with and without delayed gastric emptying, can be managed as per existing gastroparesis treatment pathways.^44,45^ On the other hand, patients with investigations suggestive of impaired accommodation could potentially benefit from accommodation enhancing medications such as buspirone.^46^ More invasive options such as pyloroplasty and G-POEM have been attempted,^47,48^ and revisional surgery to restore accommodation capacity of the proximal stomach could be attempted if symptoms are refractory.^5,49^ However, the risk of a poor symptomatic outcome increases with each repeated revisional procedure.^7,50^ Gastric resection may also be attempted as a last-resort but reported efficacy is conflicting and based on limited cases. ^51–54^ Treatment for vagal dysfunction remains a very challenging problem without effective therapies to enhance motility recovery. However, treatments for visceral hypersensitivity, and treatments targeting accommodation may be of symptomatic benefit. In patients with normal spectrograms and amplitude-independent symptoms, revisional surgery is likely to be of limited efficacy, and neuromodulating medications, psychotherapy, and a multimodal approach plausibly should be attempted as first-line therapy.^55^

Lastly, delayed gastric emptying did not differ between those with and without Gastric Alimetry spectral abnormality in this cohort. Given the inconsistent relationship between emptying time and symptoms, the lack of resolution of symptoms despite emptying improvements, and the variable repeatability over time,^56–58^ this finding may suggest that gastric emptying time offers more limited clinical utility in post-fundoplication symptoms than previously thought. Gastric emptying time has not been demonstrated to be decreased in those with post-fundoplication symptoms,^5^ and conversely may be accelerated by fundoplication.^59,60^ As such, other pathophysiological mechanisms may be of greater importance after fundoplication.

Some limitations of the study should be noted. Patient inclusion was based on an unselected consecutive sample of patients referred to two specialist gastrointestinal tertiary centres for Gastric Alimetry assessment. Prospective studies before and after fundoplication would be of interest to define whether the abnormalities identified are definitively due to the surgical procedures. Secondly, cohort size was limited, and larger prospective studies would now be desirable. Lastly, formal gastric emptying scintigraphy studies were not available for all patients. While this current study was able to highlight the prevalence of Gastric Alimetry abnormalities, future prospective studies integrating treatment algorithms in symptomatic post-fundoplication patients are needed to confirm findings. In the past, electrogastrography has not been used clinically due to significant technical limitations.^61^ However, a strength of this study is that high-resolution BSGM, using Gastric Alimetry was employed, which overcomes the limitations of EGG, and offers a new clinical opportunity to positively impact patient management and healthcare utilisation.^44^

In summary, this study has evaluated gastric myoelectrical function and symptoms using Gastric Alimetry in post-fundoplication patients, revealing distinct mechanistic phenotypes. Symptom severity was correlated with degree of myoelectrical abnormality. These findings are anticipated to improve the management of post-fundoplication patients through improved selection of patients for procedural therapies.

## Data Availability

All data produced in the present study are available upon reasonable request to the authors

## Supplementary material

**Figure S1:**
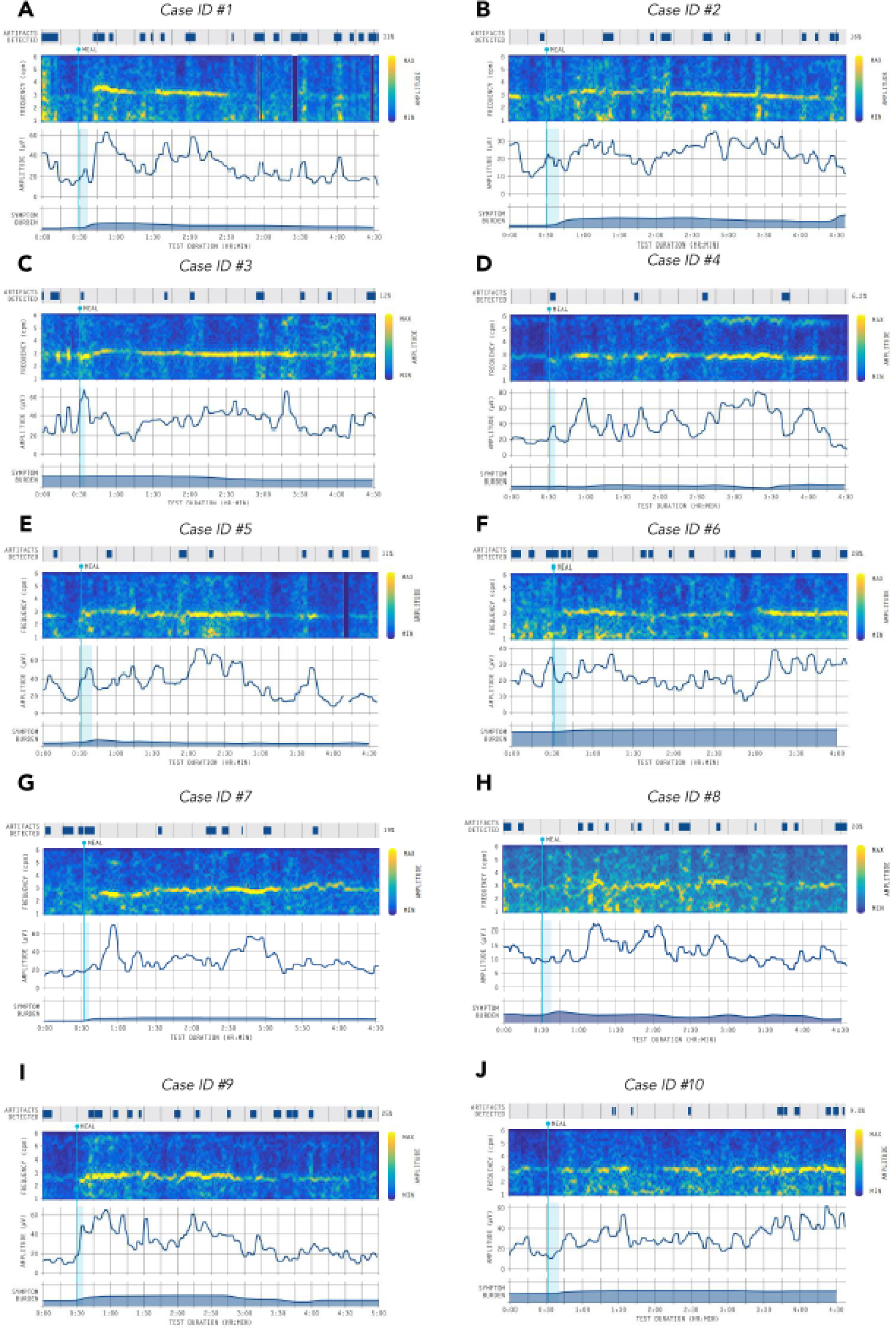
Gastric Alimetry spectral data and symptom burden plot for Cases 1-10 (A-J). Normal Alimetry Tests. Final Gastric Alimetry phenotypes ^17,20,25^ are as follows: (A-B) amplitude-independent, (C) activity-relieved, (D) post-gastric, (E-H) amplitude-independent, (I-J) sensorimotor

**Figure S2:**
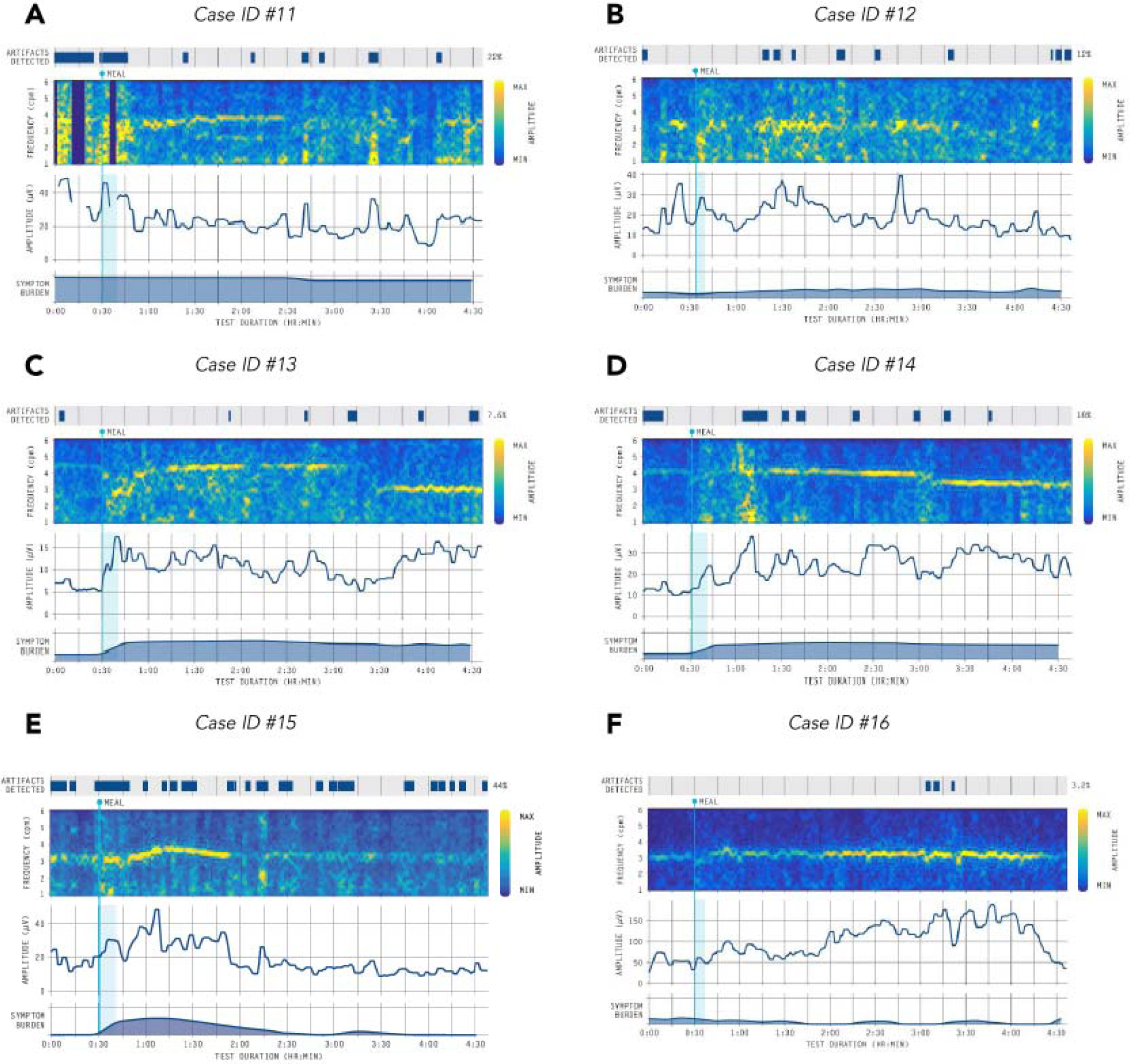
Gastric Alimetry spectral data and symptom burden plot for Cases 9-13 (A-F). Abnormal Alimetry Tests. Final Gastric Alimetry phenotypes^17,20,25^ are as follows: (A-B) low rhythm stability, (C-E) high frequency, (F) high amplitude

**Figure S3:**
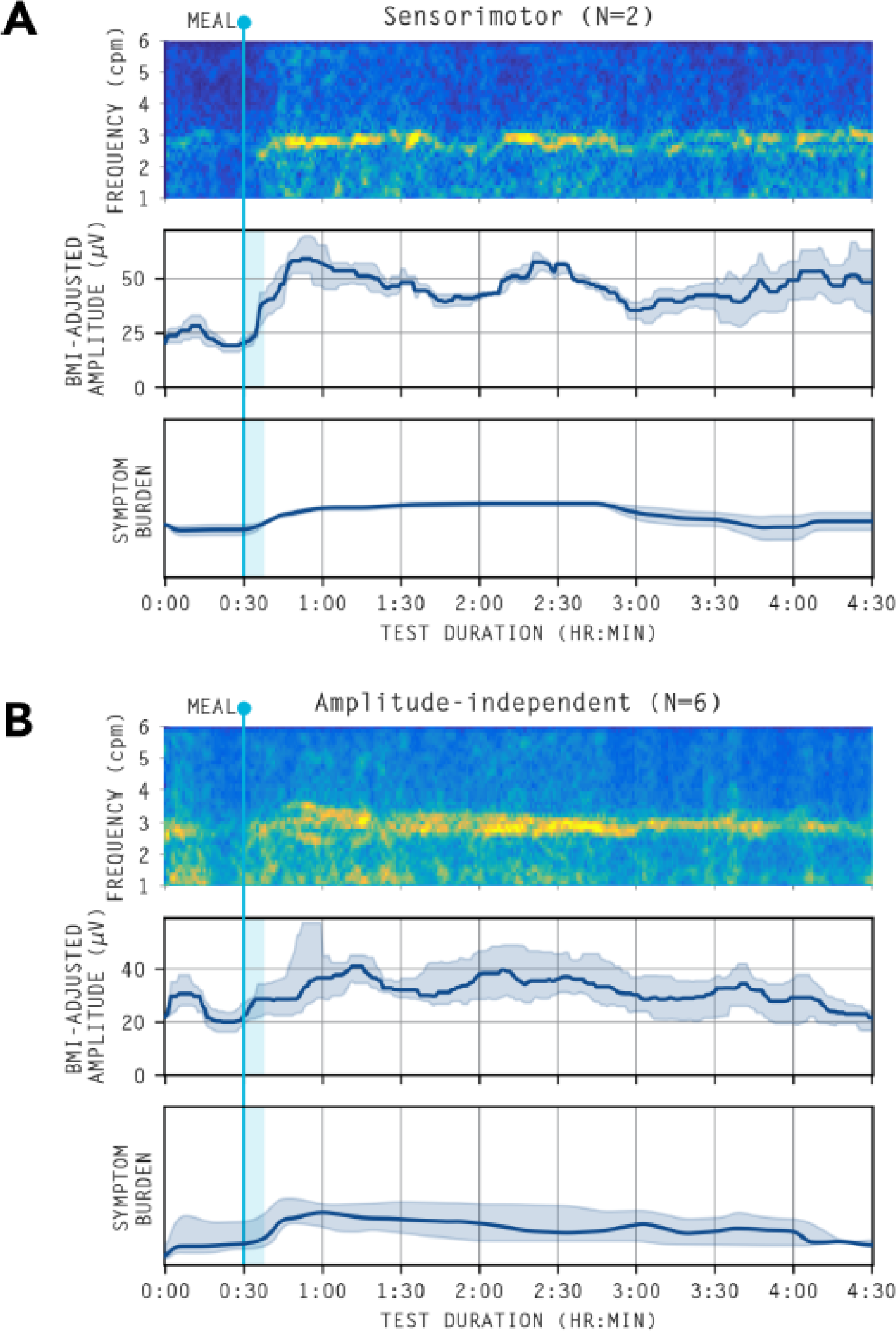
Averaged spectrograms and amplitude curves of abnormal spectrograms. (A) Sensorimotor phenotype *(amplitude-dependent*) whereby there is a high correlation between gastric amplitude and symptom burden (r>0.5). (B). *Amplitude-independent* cases whereby symptoms are continuous/do not correlate with gastric amplitude.^23^ Note that one case of a post-gastric symptom *(amplitude-dependent*) phenotype and one case of meal-relieved (*amplitude-dependent*) is not shown here but can be found in the **supplementary materials**

**Figure S4:**
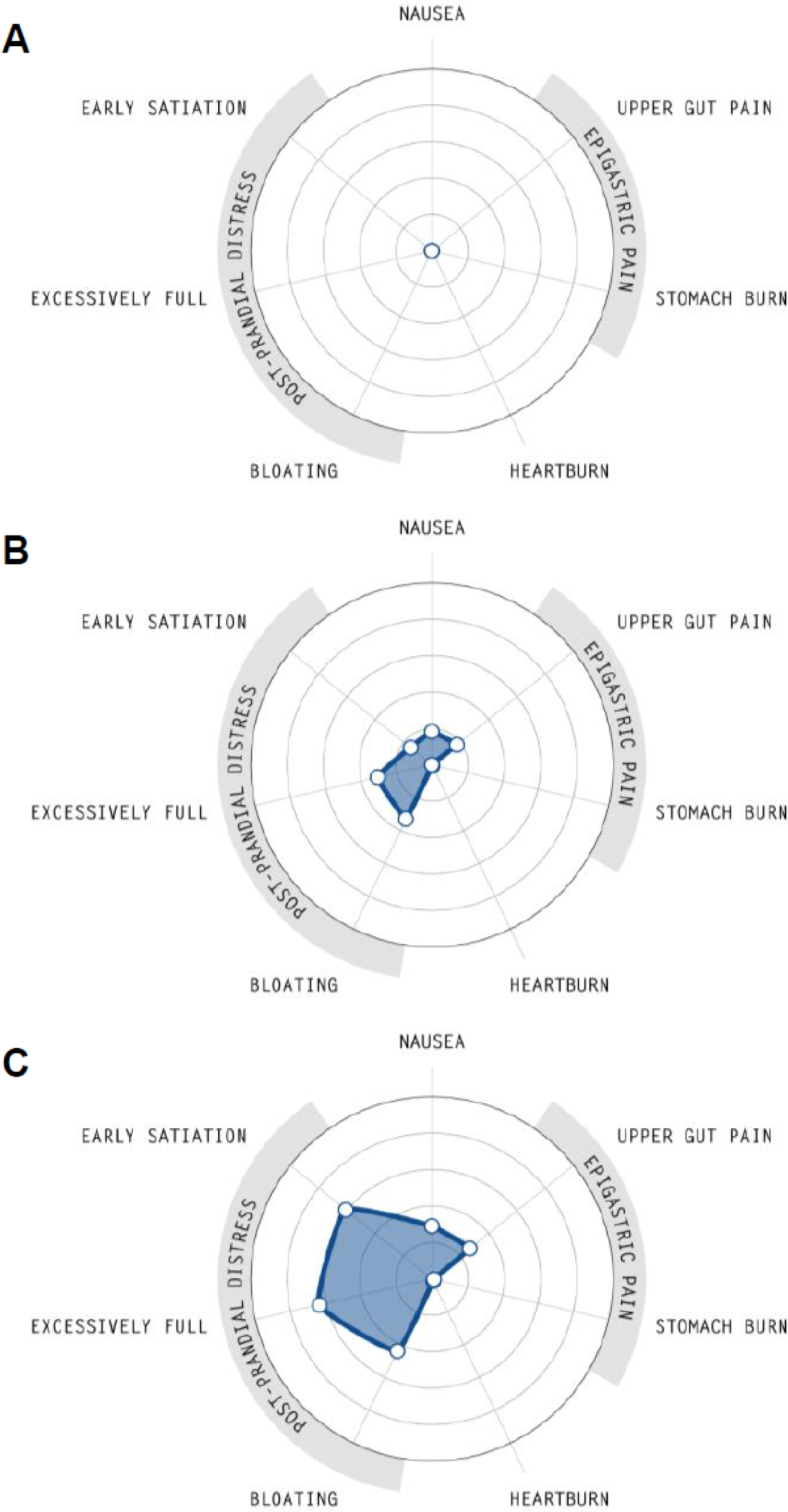
Radial plots of symptom profiles between cohorts. Median symptom characteristics between (A) controls, (B) normal and (C) abnormal Gastric Alimetry Spectral analysis.

**Table S1:**
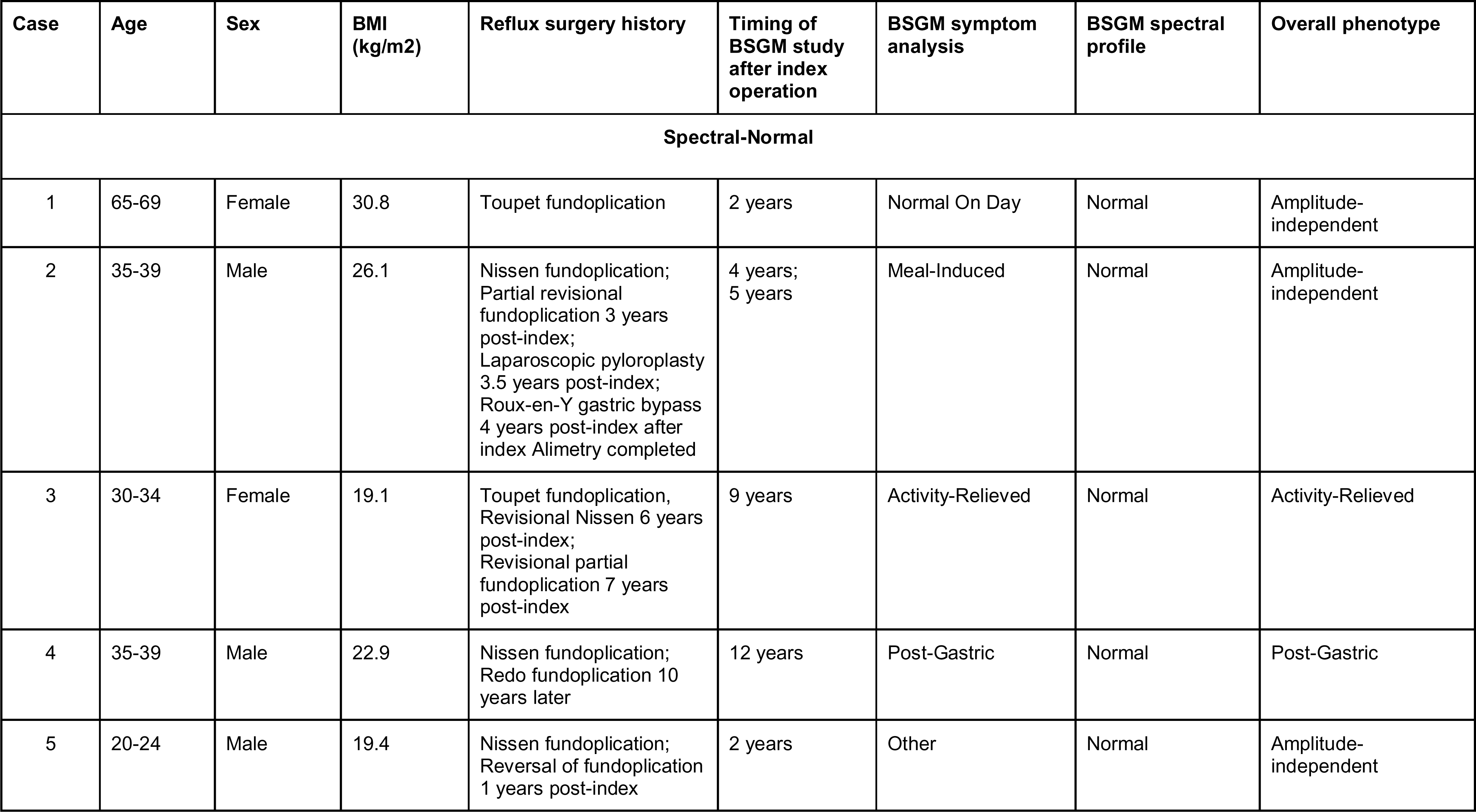
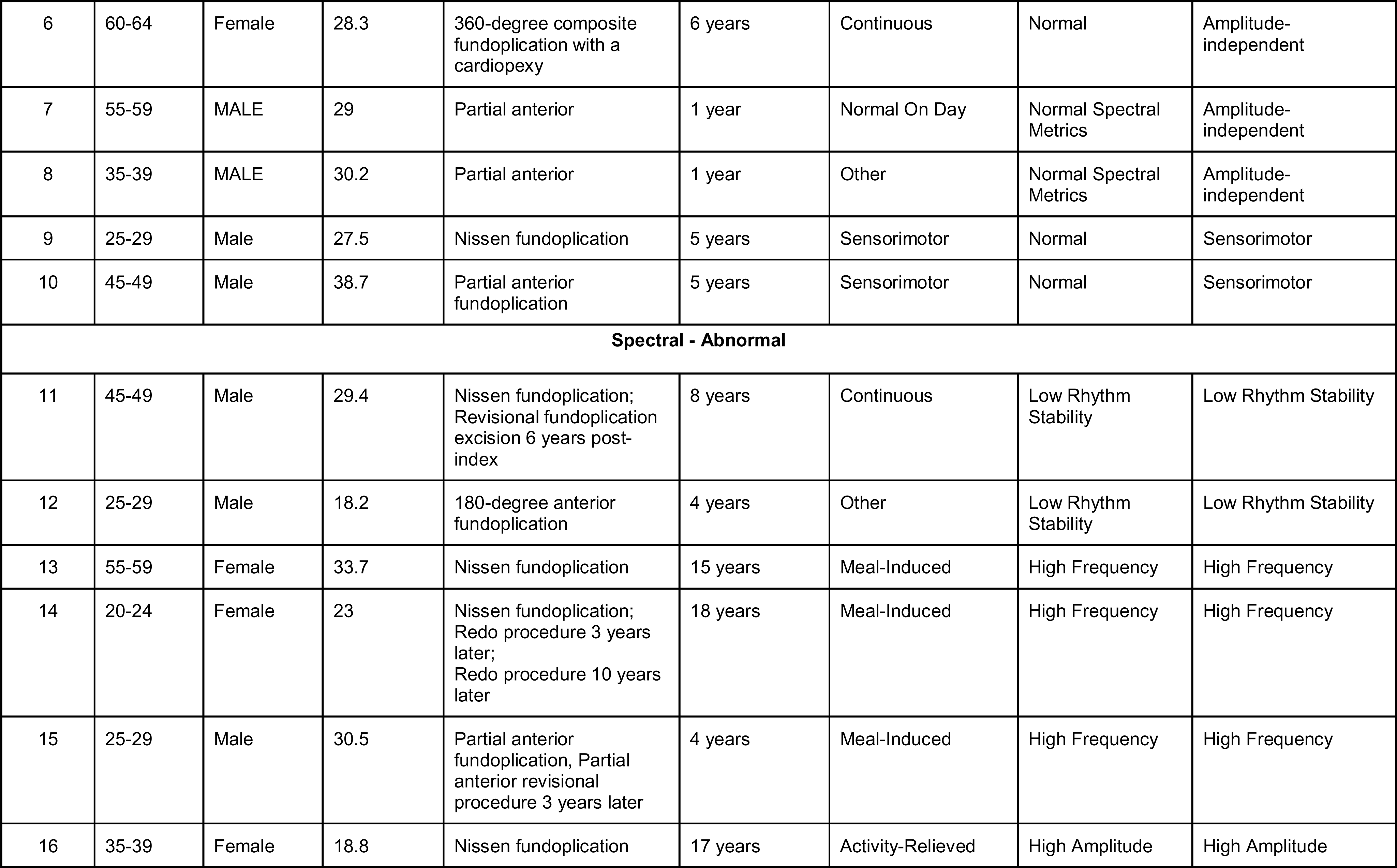
Summary of cases.

| Case | Age | Sex | BMI (kg/m<sup>2</sup>) | Reflux surgery history | Timing of BSGM study after index operation | BSGM symptom analysis | BSGM spectral profile | Overall phenotype |
| --- | --- | --- | --- | --- | --- | --- | --- | --- |
| Spectral-Normal |  |  |  |  |  |  |  |  |
| 1 | 65-69 | Female | 30.8 | Toupet fundoplication | 2 years | Normal On Day | Normal | Amplitude-independent |
| 2 | 35-39 | Male | 26.1 | Nissen fundoplication;<br>Partial revisional fundoplication 3 years post-index;<br>Laparoscopic pyloroplasty 3.5 years post-index;<br>Roux-en-Y gastric bypass 4 years post-index after index Alimetry completed | 4 years;<br>5 years | Meal-Induced | Normal | Amplitude-independent |
| 3 | 30-34 | Female | 19.1 | Toupet fundoplication, Revisional Nissen 6 years post-index;<br>Revisional partial fundoplication 7 years post-index | 9 years | Activity-Relieved | Normal | Activity-Relieved |
| 4 | 35-39 | Male | 22.9 | Nissen fundoplication;<br>Redo fundoplication 10 years later | 12 years | Post-Gastric | Normal | Post-Gastric |
| 5 | 20-24 | Male | 19.4 | Nissen fundoplication;<br>Reversal of fundoplication 1 years post-index | 2 years | Other | Normal | Amplitude-independent |
| 6 | 60-64 | Female | 28.3 | 360-degree composite fundoplication with a cardiopexy | 6 years | Continuous | Normal | Amplitude-independent |
| 7 | 55-59 | MALE | 29 | Partial anterior | 1 year | Normal On Day | Normal Spectral Metrics | Amplitude-independent |
| 8 | 35-39 | MALE | 30.2 | Partial anterior | 1 year | Other | Normal Spectral Metrics | Amplitude-independent |
| 9 | 25-29 | Male | 27.5 | Nissen fundoplication | 5 years | Sensorimotor | Normal | Sensorimotor |
| 10 | 45-49 | Male | 38.7 | Partial anterior fundoplication | 5 years | Sensorimotor | Normal | Sensorimotor |
| **Spectral - Abnormal** |  |  |  |  |  |  |  |  |
| 11 | 45-49 | Male | 29.4 | Nissen fundoplication; Revisional fundoplication excision 6 years post-index | 8 years | Continuous | Low Rhythm Stability | Low Rhythm Stability |
| 12 | 25-29 | Male | 18.2 | 180-degree anterior fundoplication | 4 years | Other | Low Rhythm Stability | Low Rhythm Stability |
| 13 | 55-59 | Female | 33.7 | Nissen fundoplication | 15 years | Meal-Induced | High Frequency | High Frequency |
| 14 | 20-24 | Female | 23 | Nissen fundoplication; Redo procedure 3 years later; Redo procedure 10 years later | 18 years | Meal-Induced | High Frequency | High Frequency |
| 15 | 25-29 | Male | 30.5 | Partial anterior fundoplication, Partial anterior revisional procedure 3 years later | 4 years | Meal-Induced | High Frequency | High Frequency |
| 16 | 35-39 | Female | 18.8 | Nissen fundoplication | 17 years | Activity-Relieved | High Amplitude | High Amplitude |

**Table S2:** Pairwise comparisons between post-fundoplication patients and controls.

| Variable | Control | Post-fundoplication | Total | p-value |
| --- | --- | --- | --- | --- |
| **Total N (%)** | **16 (50.0)** | **16 (50.0)** | **32** |  |
| Adjusted Amplitude (μV) | 37.9 (29.2 to 51.7) | 32.2 (28.9 to 40.4) | 34.5 (29.2 to 44.3) | 0.291 |
| Fed Fasted - Amplitude Ratio | 2.0 (1.2 to 2.4) | 1.8 (1.4 to 2.2) | 2.0 (1.4 to 2.2) | 0.749 |
| Principal Gastric Frequency (cpm) | 3.1 (2.9 to 3.2) | 3.0 (2.9 to 3.2) | 3.1 (2.9 to 3.2) | 0.585 |
| Principal Gastric Frequency Deviation (from 3cpm) | 0.3 (0.2 to 0.4) | 0.3 (0.2 to 0.5) | 0.3 (0.2 to 0.5) | 0.836 |
| Gastric Alimetry Rhythm Index | 0.5 (0.4 to 0.8) | 0.4 (0.3 to 0.6) | 0.5 (0.4 to 0.6) | 0.200 |
| Meal Completion (%) | 100.0 (100.0 to 100.0) | 100.0 (67.5 to 100.0) | 100.0 (97.5 to 100.0) | 0.124 |
| Percent Artefact (%) | 8.6 (4.8 to 10.1) | 17.8 (8.2 to 22.6) | 9.6 (7.0 to 19.0) | **0.016** |
| PAGI-SYM Score | 0.0 (0.0 to 0.4) | 2.6 (2.2 to 3.2) | 1.0 (0.0 to 2.6) | **<0.001** |
| GCSI Score | 0.0 (0.0 to 0.0) | 3.3 (2.4 to 3.7) | 1.3 (0.0 to 3.2) | **<0.001** |
| PAGI-QoL Score | 0.2 (0.0 to 0.3) | 2.7 (1.8 to 3.5) | 0.8 (0.2 to 2.6) | **<0.001** |
| Total Symptom Burden | 0.0 (0.0 to 0.0) | 14.8 (8.2 to 27.4) | 1.2 (0.0 to 14.6) | **<0.001** |

**Table S3:**
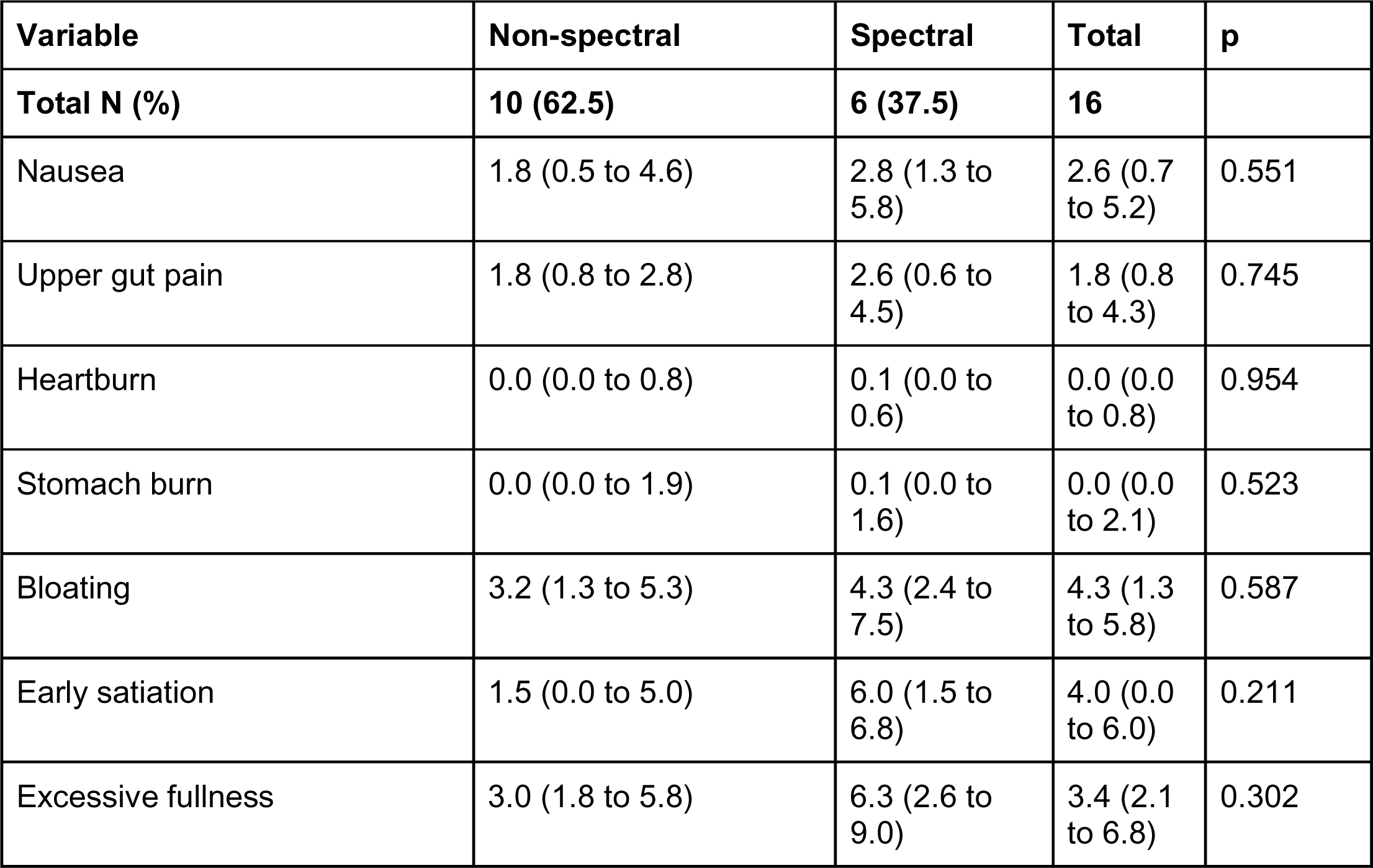
Symptom subscales for patients.

| Variable | Non-spectral | Spectral | Total | p |
| --- | --- | --- | --- | --- |
| **Total N (%)** | **10 (62.5)** | **6 (37.5)** | **16** |  |
| Nausea | 1.8 (0.5 to 4.6) | 2.8 (1.3 to 5.8) | 2.6 (0.7 to 5.2) | 0.551 |
| Upper gut pain | 1.8 (0.8 to 2.8) | 2.6 (0.6 to 4.5) | 1.8 (0.8 to 4.3) | 0.745 |
| Heartburn | 0.0 (0.0 to 0.8) | 0.1 (0.0 to 0.6) | 0.0 (0.0 to 0.8) | 0.954 |
| Stomach burn | 0.0 (0.0 to 1.9) | 0.1 (0.0 to 1.6) | 0.0 (0.0 to 2.1) | 0.523 |
| Bloating | 3.2 (1.3 to 5.3) | 4.3 (2.4 to 7.5) | 4.3 (1.3 to 5.8) | 0.587 |
| Early satiation | 1.5 (0.0 to 5.0) | 6.0 (1.5 to 6.8) | 4.0 (0.0 to 6.0) | 0.211 |
| Excessive fullness | 3.0 (1.8 to 5.8) | 6.3 (2.6 to 9.0) | 3.4 (2.1 to 6.8) | 0.302 |

